# Efficacy and safety of pre-exposure of antibiotic prophylaxis for leptospirosis: protocol for a systematic review and meta-analysis

**DOI:** 10.1101/2021.11.20.21266638

**Authors:** Benny Rashuamán-Conche, Silvana Loli-Guevara, Ethel Rodriguez-López, Carlos Alva-Díaz

## Abstract

**Introduction:** Leptospirosis is the most widespread zoonosis in the world. It represents a public health problem especially in tropical and subtropical regions, but it is also present in temperate regions. Spirochetes from *leptospira* genus cause the disease, they affect humans as an intermediate host. About pre-exposure prophylaxis for people at risk, antibiotics such as doxycycline or azithromycin were used to prevent the development of leptospirosis and its related adverse outcomes. However, the evidence about the efficacy and safety of this intervention is limited.

**Objectives:** To determine whether pre-exposure antibiotic administration prevents infection, hospitalization, or mortality from leptospirosis, without causing severe adverse effects.

**Methods:** We propose to do a systematic review and meta-analysis. We will search in Pubmed (Medline), Embase.com, Cochrane Central Register of Controlled Trials (CENTRAL), Scopus, Web of Science, LILACS and ClinicalTrials.gov. Individual randomized controlled trials, non-randomized controlled trials, cohorts, and cases-control studies will be included according to the inclusion and exclusion criteria set. The flow chart for selecting studies to be included will be presented in accordance with the PRISMA guide. The methodological quality of the studies will be evaluated by duplicate. Subsequently, the qualitative analysis of the data will be carried out and the feasibility of a quantitative meta-analysis will be evaluated. Finally, a summary of findings table will be presented according to the feasibility of the meta-analysis.

**Results:** The results will be published in a peer-reviewed journal.

**Conclusion:** This systematic review will sum up-to-date evidence about the efficacy and safety of pre-exposure antibiotic prophylaxis for preventing laboratory-confirmed leptospirosis, hospitalization and mortality.

## 1. BACKGROUND

Leptospirosis is the most widespread zoonosis worldwide and represents a public health problem especially in tropical and subtropical regions. However, it is also present in temperate regions. It is caused by spirochetes from genus Leptospira ^1^. Humans are intermediate hosts that acquire the disease through direct contact with the urine of infected animals, such as rats ^2^ or through a contaminated environment such as water ^3^.

Each year, there are about 1 million new cases of leptospirosis causing more than fifty-eight thousand deaths worldwide, while the most affected population are males between 20 and 49 years. It reaches its highest rates in the South Asia and Southeast Asia region in terms of both number of cases and deaths ^4^. Else, the underdiagnosis of leptospirosis, especially of mild forms, can mask the actual prevalence, delay treatment, increase mortality and affect disability adjusted life years ^5^.

Outbreaks of this disease usually occur after episodes of heavy rain or flooding, and have epidemic potential, but there are also behavioral factors that increase the individual risk of disease like working with livestock or swimming among others ^6,7^. Because of that, WHO emphasizes the role of occupational, recreational and social activities to categorize risk factors and risk groups for leptospirosis (such as cattle farmers, soldiers potentially exposed to contaminated water, etc.) ^8^.

Currently, different guidelines worldwide include pre-exposure prophylaxis with doxycycline or amoxicillin as an option to avoid the disease when traveling to areas of high endemicity or in groups at high risk for infection ^9–15^; however, the World Health Organization (WHO) guidance do not contemplate it ^8^. Due to the wide distribution of leptospirosis worldwide ^4^, it is important to know whether pre-exposure antibiotic prophylaxis is effective in preventing the development of leptospirosis. A previous study from 2009 that evaluated efficacy and safety of antibiotic prophylaxis to prevent leptospirosis concluded that the possible benefits with doxycycline (the only drug evaluated) were not clear with the data available until that date ^16^. In 2017, Scheneider et al. found that studies on pre-exposure antibiotic prophylaxis were inconsistent in preventing leptospira infection ^17^. In 2018, a clinical trial evaluated the efficacy of pre-exposure prophylaxis with doxycycline and azithromycin to prevent leptospira infection concluding in a reduction of seropositivity ^18^. Hence, the efficacy of pre-exposure antibiotic prophylaxis is not established to date.

The present review aims to determine whether pre-exposure antibiotic prophylaxis protects against laboratory-confirmed leptospirosis, hospitalization, or death, as well as to study its safety.

## 2. REVIEW QUESTION

Is pre-exposure antibiotic prophylaxis effective and safe?

## 3. METHODS

### Search strategy

The search will be performed on Pubmed (Medline), Embase.com, Cochrane Central Register of Controlled Trials (CENTRAL), Scopus, Web of Science, LILACS and ClinicalTrials.gov. We will not restrict studies by date, language, or publication status. For the search of adverse effects, only those reported in the studies included in the review will be considered.

We will also check the literature included in previous systematic reviews identified with the search strategies, to locate studies that meet our eligibility criteria. In addition, we will search for potential studies not identified by the search strategies by reviewing the references of the included studies, and also searching for studies that cited the included studies through Google Scholar.

Finally, we will also consider the information of comments and retractions of the included studies, to identify errata in the data or ethical misconduct that disqualify the inclusion of a study to our review.

The search strategies are attached in Appendix.

### Condition or domain being studied: Leptospirosis

#### Eligibility criteria

##### Participants/Population

Inclusion: Participants without age or gender restriction, who belong to a risk group or have risk factors for developing leptospirosis, according to the WHO guidance for diagnosis, surveillance and control ^8^.

##### Intervention

Any antibiotic regimen administered as pre-exposure prophylaxis for leptospirosis, given by any route of administration.

##### Comparators

Any comparators found in the literature: active comparator, placebo or neither.

##### Types of studies

We will include individual randomized controlled trials (RCT), non-randomized controlled trials, cohorts, and cases-control studies.

We will include non-randomized studies as the looked intervention is unlikely to be studied in randomized trials. Because of the different settings of prevalence of the disease or the lack of resources invested ^7^.

##### Context

We will include studies performed in endemic countries as well as in non-endemic areas.

### 2. Outcome

#### Primary outcomes

1. Confirmed case of leptospirosis (MAT ≥ 1:400; positive culture from blood, urine or tissue samples; leptospires in tissues demonstrated by antibodies conjugated to fluorescence markers or polymerase chain reaction or immunostaining).
2. Hospitalization from all causes.
3. Mortality from all causes.

#### Secondary outcomes

1. Adverse effects from antibiotic administration as prophylaxis for leptospirosis.
2. Hospitalization due to leptospirosis (leptospirosis confirmed by laboratory).
3. Time of illness.
4. Mortality due to leptospirosis (leptospirosis confirmed by laboratory).
5. Clinical diagnosis of leptospirosis (according to the criteria of each study included).
6. Probable case of leptospirosis (ELISA, MAT ≥ 1:100)

### Data collection and analysis

#### Study selection

The searches will be executed by the authors, exported to the RAYYAN QCRI platform (https://rayyan.qcri.org/) and the filtering of duplicate articles will be carried out. Then, the review and selection according to title and abstract will be started by two authors independently. Finally, each reviewer will read the full text versions to evaluate the fulfillment of the inclusion and exclusion criteria, determining the articles to be included in the review. In case of disagreements, they will be discussed until consensus is reached; ultimately, a third reviewer will resolve conflicts in the selection. A PRISMA flow chart will be developed to chart this process ^19^. In addition, a table of characteristics of the excluded studies will be included.

### Data extraction and management

Data extraction will be performed by two independent reviewers. The data will be collected by using a data collection sheet elaborated in Excel, the following data will be extracted:

#### Study details

- First author
- Country
- Year of publication

#### Study design

- Sample size: number of participants per group
- Type of study: RCT, non-randomized controlled trial, cohort, case-control study. Confounders analyzed: Variables processed for adjusted analysis (if applicable)

#### Characteristics of the participants

- Number of participants: in each of the groups, measured at the beginning and end of the study, rate of adherence to the assigned intervention
- Age: mean and standard deviation of each group.
- Sex: number of males in each group. Percentage of males in each group.
- Group membership or risk factor for leptospirosis presented according to WHO guidance ^8^.
- Origin: Endemic area or non-endemic area.
- Occupational risk factor: Number and frequency in each group.

#### Characteristics of the intervention

- Type of antibiotic administered, dose, frequency, duration of intervention (mean and standard deviation), route of administration, time between intervention and exposure (mean and standard deviation - mean difference)

#### Comparator characteristics

- Intervention administered, dose, frequency, route of administration

#### Characteristics of the outcomes

□ For the qualitative variables, the number and proportion of participants with each of the outcomes studied will be collected.
  □ Confirmed diagnosis of leptospirosis and method used: MAT, culture, PCR, IF)
  □ Probable case of leptospirosis (ELISA, MAT ≥ 1:100)
  □ Clinical diagnosis of leptospirosis
  □ Mortality due to leptospirosis
  □ Mortality from all causes
  □ Hospitalization for leptospirosis
  □ Hospitalization for all causes
  □ Any adverse effects.
□ For quantitative variables, we will collect the mean and standard deviation
  □ Time of illness.
  □ Follow-up time until the measurement of the outcomes.
□ Type of analysis performed for each outcome (protocol or intention to treat for RCT)
□ The effect measures will be collected as raw and adjusted aggregate measures.
□ Other outcomes studied

The authors will analyze the existence of data errors or unethical research behavior in the selected articles before their inclusion in the analysis. If there are corrections that invalidate the results presented, these studies will be eliminated, in order to maintain the quality of the data included.

### Risk of bias assessment

Two authors will conduct the risk of bias assessment of each included study independently. Disagreements will be resolved by discussion between the two authors until consensus is reached; if disagreement persists, it will be resolved by a third author.

The criteria to be evaluated for the risk of bias of individual randomized clinical trials will be those of Cochrane’s “RoB 2” tool for each outcome ^20^. Assessment of risk of bias for non-randomized studies will be performed using the ROBINS-I tool ^21^.

These domains will be summarized in an overall risk of bias categorized as “low risk”, “some concerns” or “high risk of bias” for randomized controlled trials ^20^. For non-randomized studies, the overall risk of bias will be categorized as “low”, “moderate”, “serious” or “critical” ^21^. This will be presented in a specific table to show the risk of bias of each included study.

#### Unit of analysis issues

If clinical trials with more than one intervention arm are found, it will be verified whether these arms meet the eligibility criteria for inclusion in the review. If this is the case, the intervention group will be considered for the general meta-analysis as the sum of the intervention arms (number of participants and events) counting the control group only once for both arms. For subgroup comparisons, the data of the control group will be divided approximately equally between the intervention arms.

#### Dealing with missing data

If necessary, data not available to the authors of the corresponding articles will be requested by contacting the corresponding author by e-mail.

In case of failure to obtain the missing data, the researchers will discuss whether the missing data from the studies were omitted randomly or not. If data are considered not missing at random, the analysis will require imputations. We will impute the missing data with the mean and treat these as if they were observed such in the Handbook Cochrane is recommended ^22^.

#### Assessment of heterogeneity

It will be evaluated according to the Cochrane methodology ^22^:

a. The evaluation of the overlap of the confidence intervals in the forest plot.
b. The application of the Chi^2^ test with a statistical significance level of 10%.
c. The application of the I^2^ test with the following interpretation:
  i. 0%-40%: May not be important
  ii. 30%-60%: May represent moderate heterogeneity
  iii. 50%-90%: May represent substantial heterogeneity
  iv. 75%-100%: Considerable heterogeneity

Heterogeneity will be identified on the basis of the above considerations. The causes of heterogeneity will be investigated through subgroup analysis.

### Data synthesis

The qualitative analysis will be carried out by means of a synthesis of the evidence found, in addition we will present the most important data of the studies in a summary table.

We will carry out a meta-analysis for each outcome with at least two studies with ‘not important’ to ‘moderate’ heterogeneity (I^2^ ≤ 60%), plus a low or low/moderate risk of bias (for RCTs and other study designs respectively). All the included studies which meet above conditions will be part of the main meta-analysis, regarding of study designs.

The quantitative analysis of the studies will be done with R language. The odds ratio will be chosen as the summary measure of the effect for dichotomous variables in case of including at least one case-control study for the meta-analysis. Otherwise, the relative risk will be preferred, which will be taken for RCTs as unadjusted data reported in the article or will be calculated if the included studies provide sufficient data for its calculation. For non-randomized studies, adjusted effect estimates will be selected by authors in order to attempt to minimize the risk of bias due to confounding. If neither of the two measures of effect can be collected, the measure that has been used in most of the included studies will be used. Likewise, the 95% confidence intervals will be presented. The summary measure of effect for continuous variables will be the mean difference.

We will adopt a fixed-effect model (by Mantel-Haenszel method) if I^2^ is less or equal to 40% and a random-effects model if it is greater than 40%; and these results will be presented in a forest plot. If meta-analysis will not be carried out; the included studies will be presented in a forest plot (including I^2^) but without a pooled estimate.

### Publication bias

We will use a funnel plot to present effect sizes plotted against their standard errors or precisions (the inverse of standard errors) **for main outcomes**. Symmetry of the plot will be assessed by this funnel plot if at least 10 studies are included. Aside, we will use Egger’s test (for funnel plot asymmetry) to regress the standardized effect sizes on their precisions; in the absence of publication bias, the regression intercept is expected to be zero ^22,23^.

### Subgroup analysis

Analysis will be conducted by subgroup according to age (over or under 18 years old); by subgroup according to the antibiotic used for prophylaxis; by population origin (endemic or traveling area); and by occupation or risk factor involved in acquiring leptospirosis as found in the included studies.

If differences are found between overall and subgroup results, related effect modifiers will be discussed. Metaregression will be applied to determine whether or not these modifications are significant.

### Sensitivity analysis

Sensitivity analysis will be performed for each primary outcome. We will perform a meta-analysis with only previously included RCTs (low risk of bias) and other with only non-randomized studies (low/moderate risk of bias).

An additional sensitivity analysis for the primary outcomes without the study with the largest sample will be added too.

### Certainty of evidence and summary of findings table

The quality of evidence will be evaluated according to the GRADE methodology, which considers 5 recommended domains (risk of bias, inconsistency, imprecision, indirectness, and publication bias) for each outcome and will be presented in the summary of findings table. For these judgments, we will assess evidence of the studies which contribute data to the meta-analysis. The certainty of the evidence ratings will be high, moderate, low, or very low ^24,25^.

### Language

English

### Limitations

Leptospirosis is a neglected disease and, consequently, we suspect there are few studies that evaluate the topic of interest.

## Supporting information

SEARCH STRATEGY PUBMED

## Data Availability

All data produced in the present study are available upon reasonable request to the authors

