## Supplementary material for "Efficacy and safety of pre-exposure of antibiotic prophylaxis for leptospirosis: protocol for a systematic review and meta-analysis": SEARCH STRATEGY PUBMED

| Search Strategy PUBMED | | | |
|---|---|---|---|
| Population | | - | |
| Concept | 1 | Prophylaxis | ("Antibiotic Prophylaxis"[Mesh] OR "prophyla*"[tiab] OR “prevention and control”[Subheading] OR prevent*[tiab] OR "Pre-Exposure Prophylaxis"[Mesh] OR "chemoprophyla*"[tiab] OR “chemoprevent*”[tiab]) |
| | 2 | Antibiotics | ("Anti-Bacterial Agents"[Mesh] OR "Doxycycline"[Mesh] OR "Azithromycin"[Mesh] OR "Penicillins"[Mesh] OR "doxycycline"[tiab] OR "azithromycin"[tiab] OR penicillin*[tiab] OR "Amoxicillin"[tiab] OR "Antibiotic Prophylaxis"[Mesh] OR "antibiotic*"[tiab] OR "antibacterial*"[tiab] OR "anti-bacterial*"[tiab]) |
| | 3 | Leptospirosis | ("Leptospirosis"[Mesh] OR “Leptospira”[Mesh] OR ("leptospir*"[tiab] OR "leptosporosis"[tiab] OR "Stuttgart Disease*"[tiab] OR "Mud Fever"[tiab] OR "Rice-Field Fever"[tiab] OR "Rice Field Fever"[tiab] OR "Cane-Cutter Fever"[tiab] OR "Cane Cutter Fever"[tiab] OR "Swineherd's Disease*"[tiab] OR "Canicola fever"[tiab] OR “Weil Disease”[tiab] OR “Weils Disease”[tiab] OR “Weil Syndrome”[tiab] OR "Spirochetal Jaundice"[tiab]) OR ("leptospir*"[ot] OR "leptosporosis"[ot] OR "Stuttgart Disease*"[ot] OR "Mud Fever"[ot] OR "Fever, Mud"[ot] OR "Rice-Field Fever"[ot] OR "Fever, Rice-Field"[Ot] OR "Rice Field Fever"[ot] OR "Cane-Cutter Fever"[ot] OR "Cane Cutter Fever"[ot] OR "Fevers, Cane-Cutter"[ot] OR "Swineherd's Disease*"[ot] OR "Canicola fever"[ot] OR "Fever, Canicola"[ot] OR “Weil Disease”[ot] OR "Disease, Weil"[ot] OR "Weil's Disease"[ot] OR "Disease, Weil's"[ot] OR "Weils Disease"[ot] OR “Weil Syndrome”[ot] OR "Jaundice, Spirochetal"[ot] OR "Spirochetal Jaundice"[ot])) |
| Study | 4 | RCT | (((randomized controlled trial[pt]) OR (controlled clinical trial[pt]) OR (randomized[tiab] OR randomised[tiab]) OR (placebo[tiab]) OR (drug therapy[sh]) OR (randomly[tiab]) OR (trial[tiab]) OR (groups[tiab])) NOT (animals[mh] NOT humans[mh])) |
| | 5 | Non-randomized studies | (cohort[all] OR (control[all] AND study[all]) OR (control[tw] AND group*[tw]) OR epidemiologic studies[mh] OR program[tw] OR clinical trial[pt] OR comparative stud*[all] OR evaluation studies[all] OR statistics as topic[mh] OR survey*[tw] OR follow-up*[all] OR time factors[all] OR ci[tw]) NOT ((animals[mh:noexp] NOT humans[mh:noexp]) OR comment[pt] OR editorial[pt] OR review[pt] OR meta analysis[pt] OR case report[tw] OR consensus[mh] OR guideline[pt] OR history[sh]) |
| Final | ((1 **AND** 2) **AND** 3) **AND** (4 OR 5) | | |
